# Autism Research at a Crossroads: Global Progress, Persistent Gaps, and Future Pathways : A Bibliometric Analysis

**DOI:** 10.64898/2026.07.14.26358066

**Authors:** Qianyi Zhong, Luping Chen, Yue Ji, Fenglei Zhu, Xiaobing Zou

**Affiliations:** Child Development Behavior Center, The Third Affiliated Hospital of Sun Yat-sen University, Guangzhou 510630, China; Cerebrovascular Surgery, The Third Affiliated Hospital of Sun Yat-sen University, Guangzhou 510630, China

**Keywords:** Autism spectrum disorder, bibliometrics, China, data visualization, research trends

## Abstract

**Background:** The global prevalence of autism spectrum disorder (ASD) has significantly increased over the past two decades. Despite substantial research advances, critical aspects, including etiology, diagnostic biomarkers, and pharmacological interventions, remain incompletely elucidated. This persistent knowledge gap warrants systematic mapping of the field’s evolution to inform future research priorities.

**Methods:** A bibliometric analysis of ASD-related publications indexed in Web of Science was conducted from January 2020 to May 2025. Following a systematic deduplication process, original articles, reviews, case reports, and clinical trials were included in the analysis. The analytical framework comprised co-authorship networks, institutional collaboration patterns, national research contributions, and keyword co-occurrence structures, all of which were examined using CiteSpace (version 5.8.R3) and VOSviewer.

**Results:** After deduplication, 8,162 publications (January 2020–May 2025) were analyzed. The annual output grew steadily, confirming ASD as a sustained priority in neuroscience. Research remains academia-driven, led by the United States, with China as the second-largest contributor. Chinese institutions place greater emphasis on mechanistic and developmental phenotyping, which aligns with national priorities. These studies maintain strong methodological rigor, and their growing volume underscores the central role of ASD in translational neuroscience.

**Conclusion:** Future research on ASD should focus on strengthening case identification, refining clinical phenotyping, and expanding large-scale cohort studies to advance our understanding of its etiology and identify reliable diagnostic biomarkers. It is equally important to develop and evaluate targeted interventions for core symptoms and integrate telemedicine into service delivery models. A critical yet understudied priority is improving the quality of life for autistic individuals and their families, an area in which research globally, including in China, requires greater depth and consistency. With China’s growing investment in autism research, it is well-positioned to contribute to these pressing international challenges.

## 1. Introduction

Autism spectrum disorder (ASD) is a neurodevelopmental disorder marked by early onset, chronic progression, and significant heterogeneity [1], with core symptoms encompassing impaired social communication, restricted interests, and repetitive behaviors. Since the 1990s, the reported prevalence of ASD has increased from approximately 0.7% to 1.0% [1, 2]. Recent data, including a 2018 Centers for Disease Control and Prevention (CDC) study reporting that 1 in 44 eight-year-old children in the United States are affected, suggest this upward trend continues [3]. This increase may partly reflect heightened awareness, broadened diagnostic criteria, and recognition of a wider phenotypic spectrum [4,5]. ASD exhibits a gender disparity, with a male-to-female ratio of 3-4:1 [6], and is frequently associated with co-occurring conditions, such as attention-deficit/hyperactivity disorder, intellectual disability, and sleep disorders [7,8], thereby representing a leading cause of intellectual disability in young children [9]. Currently, there are no specific diagnostic biomarkers [10] or targeted therapies [11] for ASD, resulting in substantial burdens on individuals, families, and society[12]. From a healthcare delivery perspective, the rising prevalence of ASD translates into growing demands for primary care, mental health services, rehabilitation, and long-term support systems. However, access to evidence-based early intervention remains highly uneven across regions and socioeconomic groups, even within high-income countries [13]. Diagnostic delays are common, with many children receiving a confirmed diagnosis only after entering school, thereby missing the critical window for early intervention[14]. In low- and middle-income countries, including large parts of China, shortages of trained child psychiatrists, standardized screening tools, and family-centered services further exacerbate these disparities [15]. Moreover, the quality of life (QoL) of autistic individuals and their families, recognized as a core outcome by the 2021 Lancet Commission on the future of care and clinical research in autism, has received far less research attention than biological or diagnostic investigations [16]. Improving QoL requires clinical interventions and accessible community support, educational inclusion, vocational training, and mental health services for caregivers [17].

Despite its growing prevalence, ASD remains a scientifically pressing issue, particularly in the field of etiological research [18]. Future studies should prioritize enhancing case identification, refining clinical phenotyping, expanding large-scale cohorts, and advancing etiological and biomarker research while incorporating randomized controlled trials (RCTs) for core symptoms and telemedicine approaches [19]. Crucially, improving the QoL of autistic individuals and their families is an urgent yet under-addressed goal globally, including in China. Given China’s increasing investment in autism research, it is well-positioned to contribute meaningfully to these international challenges [20]. However, translating research into clinical practice requires addressing several domestic barriers in China, including uneven distribution of diagnostic resources between urban and rural areas, low rates of early screening in primary care, limited availability of parent-mediated interventions, and insufficient large-scale, longitudinal studies that capture real-world outcomes [21]. Therefore, understanding China’s unique research trajectory, from mechanistic studies to emerging interests in telemedicine and scalable service models, is essential for global efforts to reduce disparities in ASD healthcare. Bibliometric analysis offers a powerful and reproducible method to systematically map the evolution of a research field, identify knowledge gaps, and guide future funding and policy decisions [22]. Unlike narrative reviews, bibliometric approaches quantify publication trends, collaborative networks, keyword hotspots, and citation impact. Several bibliometric studies have been conducted on ASD, focusing on areas such as genetics, neuroimaging, gut microbiota, or pharmacological treatments. Nevertheless, most existing studies are limited by short time windows, single-database searches, or a purely Western perspective [23]. To date, no bibliometric study has adopted a dual-perspective (global versus China) framework that explicitly links research outputs to healthcare priorities, such as early diagnosis, access to intervention, telemedicine implementation, and QoL. To objectively assess the current landscape and guide future directions, we conducted a bibliometric analysis of ASD-related literature from Web of Science between January 2020 and May 2025 using CiteSpace and VOSviewer [24]. We examined publication trends, key contributors, citation networks, keyword evolution, and research focus areas, such as etiology and diagnostic markers [25], aiming to provide a comprehensive overview that addresses the limitations of prior bibliometric studies and supports the development of more effective research and intervention strategies. Specifically, this study seeks to (1) depict global publication trends and collaborative structures; (2) identify major research themes, emerging hotspots, and persistent clinical gaps; (3) characterize China’s position and distinctive research characteristics within the global ASD research landscape; (4) highlight underfunded areas with high clinical relevance, especially QoL, telemedicine, and family-centered implementation science. Accordingly, we provide an evidence-based roadmap for researchers, clinicians, and policymakers to prioritize future efforts that improve outcomes for autistic individuals and their families.

## 2. Materials and Methods

### 2.1 Search strategy

A bibliometric analysis of ASD-related publications indexed in the Web of Science Core Collection was conducted, covering the period from January 2020 to May 2025. This timeframe was deliberately selected for two reasons: first, the COVID-19 pandemic has profoundly reshaped global research collaboration and output patterns, and initiating the analysis in 2020 allows for the capture of the latest dynamics and emerging trends in the post-pandemic era; second, a focused five-year window ensures that the findings offer timely and actionable insights for current and near-future research planning. Following a systematic deduplication process, original articles, reviews, case reports, and clinical trials were retained for further evaluation. The analytical framework comprised co-authorship networks, institutional collaboration patterns, national research contributions, and keyword co-occurrence structures, all of which were examined using CiteSpace (version 5.8.R3) and VOSviewer.

Article data were retrieved from Web of Science Core Collection Citation (Science Index Expanded database) , followed by a standardized screening process. The search strategy was developed using equivalent search terms across database, including “ autism” OR “ autism spectrum” OR “ autism spectrum disorder” OR “ Asperger syndrome.” The search period was from January 1, 2020, to May 31, 2025. After merging the records from the two databases, duplicates were removed. Only publications classified as articles, reviews, case reports, and clinical trials were included in the subsequent analysis. Bibliometric analysis and visualization were performed using CiteSpace (version 5.8.R3). The parameters were configured as follows: linkage retention factor=3, lookback years=5, years per slice=3, selection criteria=top 50, and pruning method=pathfinder, with pruning applied to sliced and merged networks. The overall workflow of this bibliometric study is summarized in Fig. 1.

**Fig. 1.**
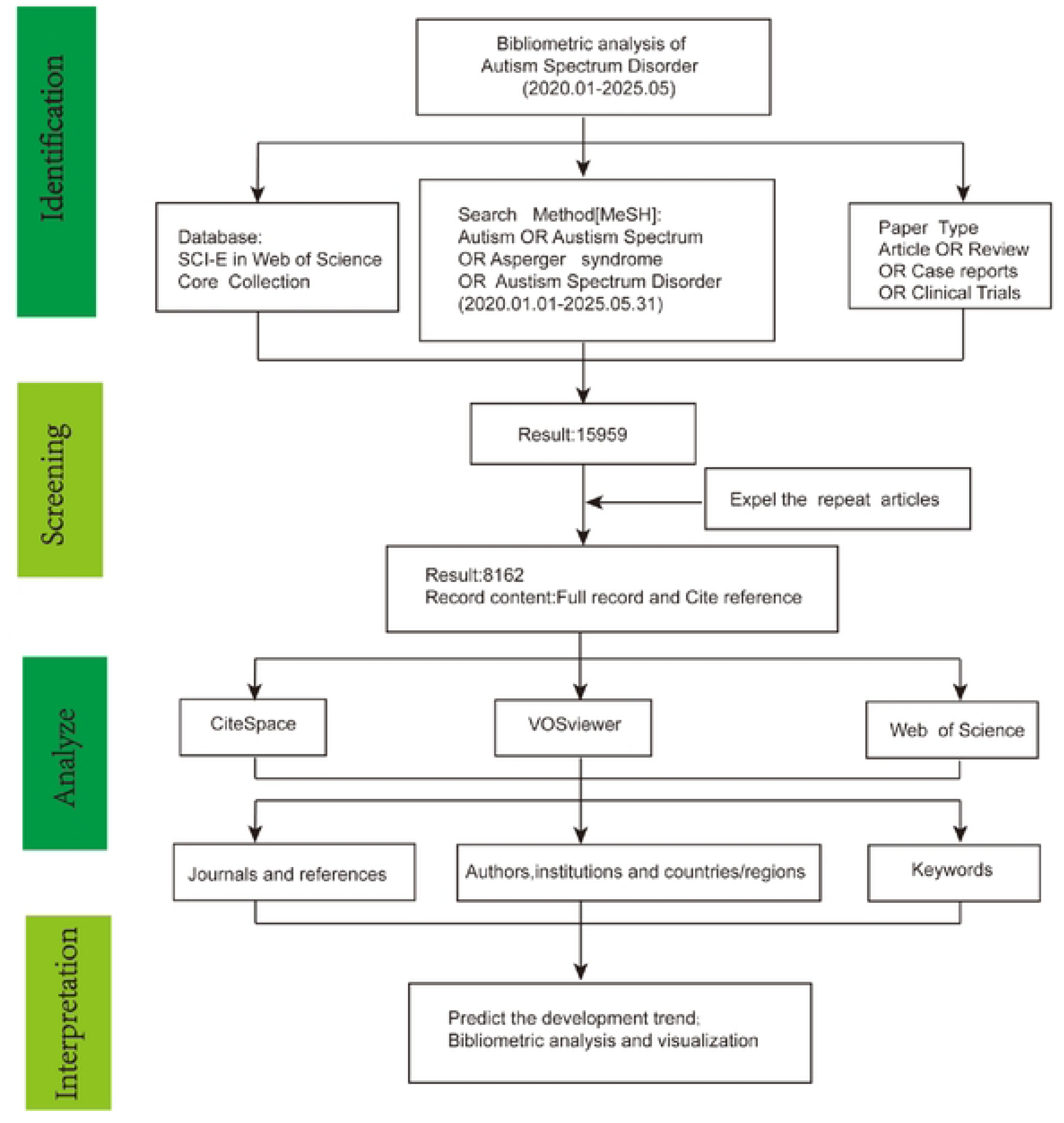
Flowchart of bibliomebic analysis.

### 2.2 Statistical analysis

This study employed bibliometric methods and utilized three specialized software tools for data analysis. First, the bibliometrix package in R was used for preliminary data processing and basic bibliometric indicator statistics. Second, CiteSpace software was applied for keyword clustering, timeline analysis, and centrality calculations for countries, institutions, authors, and keywords. This tool facilitates scientometric and data-visualization analyses and intuitively presents the structure, patterns, and distribution of disciplinary knowledge, thereby revealing the evolutionary trajectory and research frontiers of the field [22]. Additionally, VOSviewer was used to construct and visualize bibliometric networks, including relationships such as co-citation, bibliographic coupling, and collaboration, to further elucidate connections and structures among studies. VOSviewer was employed to analyze the clustering of countries, institutions, authors, journals, and keywords, along with metrics such as publication counts, citations, and link strength for each element [25]. In the initial search, 15,959 publications were identified, including research articles (n=8,693), reviews (n=1,769), case reports (n=577), clinical trials (n=110), and other document types (n=5,747). After duplicate removal and screening of publications from 2020 to 2025, 8,162 publications were retained for final analysis. To maintain thematic relevance in the visualization, only nodes directly related to ASD were displayed. Additionally, a minimum threshold of three was set for node inclusion; thus, only elements (countries and keywords) that met this criterion appeared on the maps. In the generated networks, link width indicates the strength of collaboration between nodes, while node size corresponds to citation volume.

### 2.3 Patient and Public Involvement

No patients or members of the public were involved in the design, conduct, analysis, or dissemination of this bibliometric study, as it is based entirely on publicly available published literature. Therefore, specific patient or public involvement activities were not applicable.

## 3. Results

### 3.1 Annual publishing trends

Annual publications on autism have consistently exceeded 1,000 articles over the past five years, exhibiting a near-continuous upward trend (Fig. 2). The peak occurred in 2021, with 1,710 publications, reflecting an annual growth rate of 15%-20%. Fig. 2A reveals that publications from 2020 received the highest citation counts, with 342 papers collectively accumulating more than 1,368 citations. Fig. 2B lists the top 10 contributing authors, whose citation frequency has risen steadily over time. The most prolific journal was The Journal of Autism and Developmental Disorders from the United States (Fig. 2C). Overall, relevant articles appeared in 340 different journals; the top 15 journals by volume are ranked in Fig. 2D. The Journal of Autism and Developmental Disorders led the article output, followed by Autism Research, Frontiers in Psychiatry, and Autism. These journals have published over 20 clinical trials in the past five years, with a steadily increasing annual output, indicating consistent publication activity in this field.

**Fig. 2.**
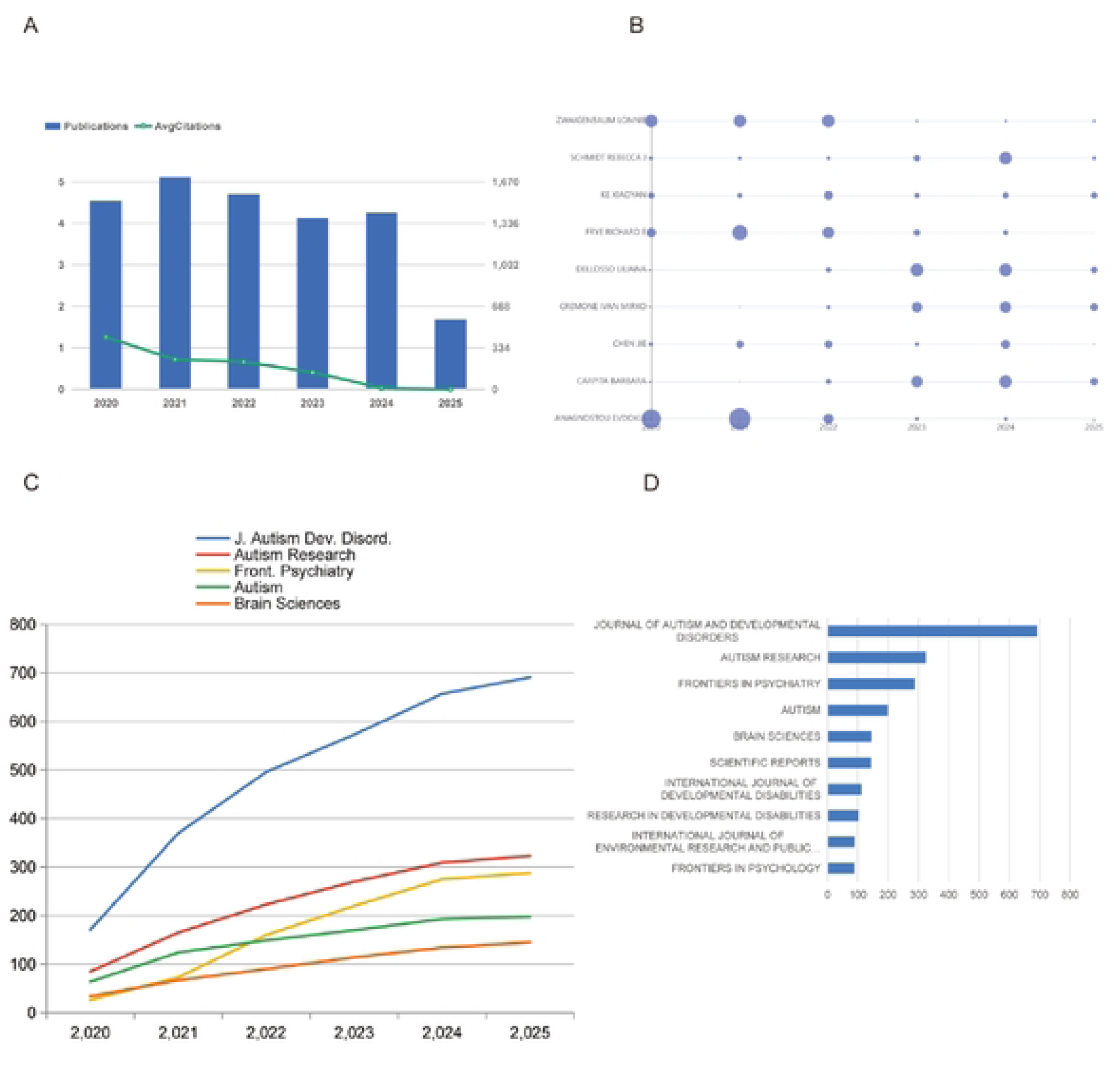
Key bibliometric trends in ASD research from 2020 to 2025. (A) Annual publication output and average citation count per year. (B) Distribution of articles based on corresponding authors’ countries. (C) Temporal trends in publications from major contributing institutions. (D) Publication volume of the top10 most productive journals in this field.

### 3.2 International Collaboration Networks in ASD Research

Fig. 3 presents an international collaboration network in autism research. Figs. 3A and B indicate that the United States leads in collaboration networks, followed by the United Kingdom, Germany, Denmark, and Australia. Although China ranks second in terms of total publication volume, its level of international engagement remains comparatively limited. European and American countries exhibit higher Multiple Country Publication (MCP) ratios, typically ranging from 30% to 40% (Fig. 3C). In contrast, the single-country publication (SCP) ratio in China exceeds 50%, suggesting greater reliance on domestic collaborations. The collaboration map illustrates global partnership patterns, revealing sparse international networks and relatively limited international connectivity. In the figure, color intensity corresponds to a country’s publication count, while the thickness of connecting lines represents the strength of collaboration between countries.

**Fig. 3.**
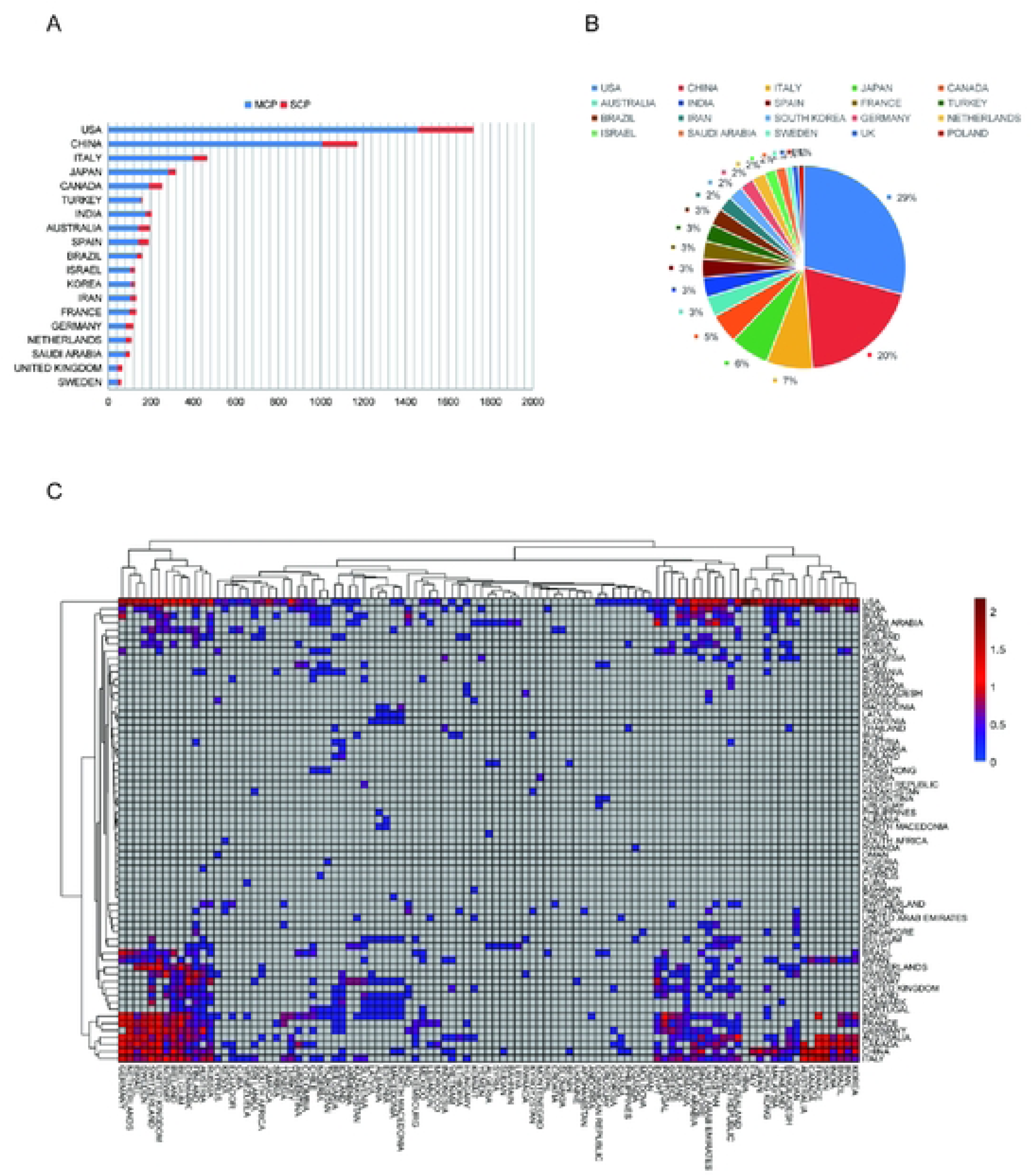
Global collaborative network in ASD research.(A)The 20 most productive countries over the *past* five years, with countries having co-corresponding authors highlighted in blue. (B) Worldwide geographical distribution of ASD research contributions. (C) International scientific collaboration relationships among countries, illustrating the strength and patterns of cooperation, with link weights presented on a base-10 logarithmic (lgl0)scale.

### 3.3 Co-authorship, Citation, and Institutional Networks in ASD Research

The co-authorship network of the 10 most prolific authors in ASD research revealed key collaborative structures (Fig. 4A). Links represent collaborative ties, and node colors indicate the activity timeline (cooler tones for earlier years and warmer tones for recent years). Nodes encircled by a purple ring denote authors with high centrality, indicating relatively high network connectivity. The two most productive contributors are affiliated with the University of Alberta and the University of Toronto, both leading Canadian institutions. Fig. 4B presents an author co-citation analysis in ASD research, where larger font sizes correspond to higher citation frequencies. Among the most-cited references, the study by Maenner MJ (2002) ranked highest. At the institutional level, Fig. 4C indicates that international institutions dominate collaborative ASD research. Although one Chinese scholar ranks tenth globally, large-scale international collaborations involving Chinese institutions remain limited. Despite China ’ s rapid growth in total publication output, its proportion of co-publications with leading international partners remains significantly lower than that of other major research regions, with most studies still conducted primarily within domestic teams.

**Fig. 4.**
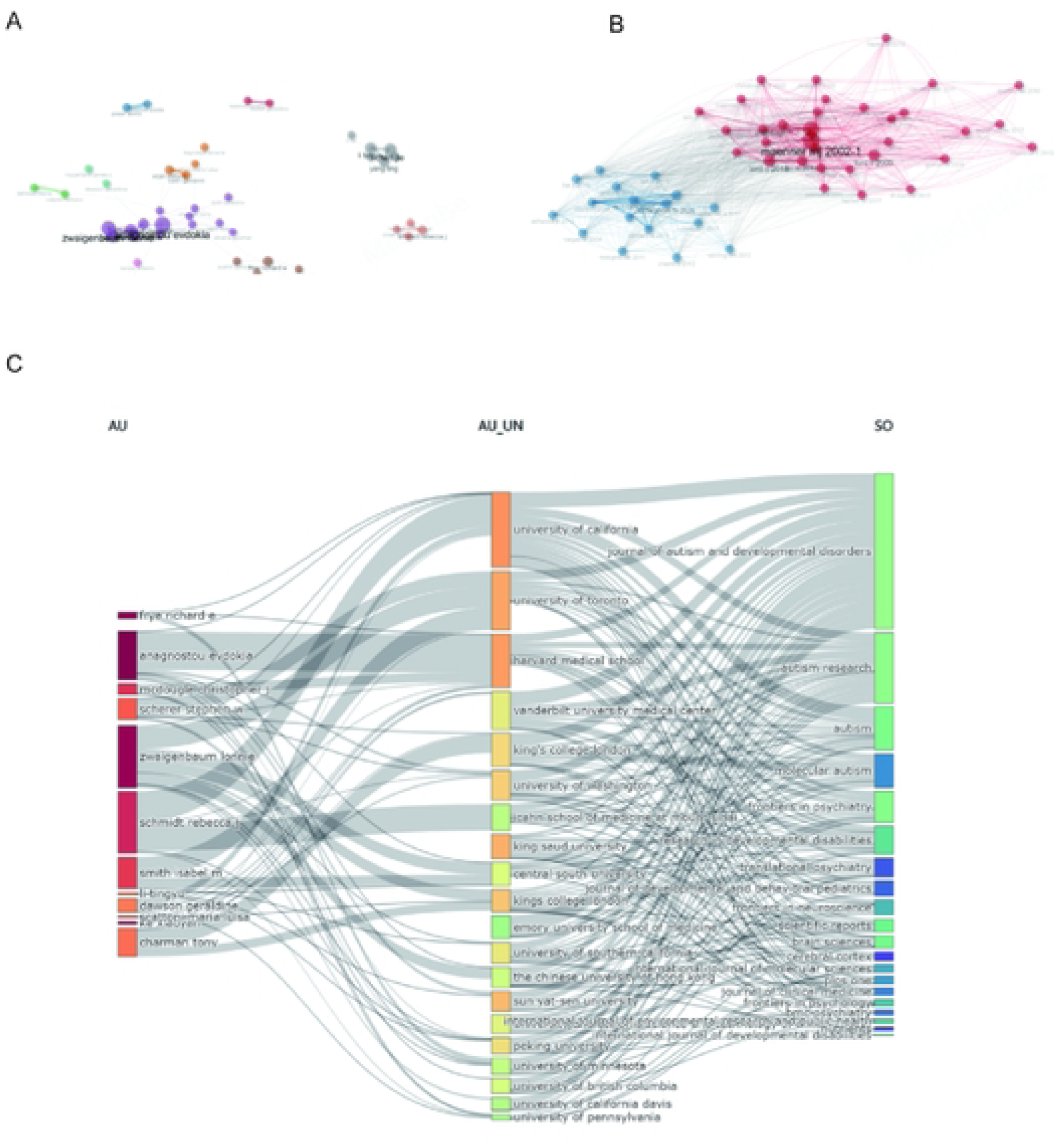
Analysis of collaboration and knowledge networks in ASD research. (A) Co-authorship network mapping collaborative relationships among researchers. (B) Co-citation network of publications reveals thematic clusters and intellectual foundations. (C)Collaborative linkages among the top10 contributing authors, institutions, and journals, providing a multi-level perspective on the field’s social structure.

### 3.4 Keywords analysis in ASD

Fig. 5A illustrates the multifactorial etiology of autism, encompassing genetic, neurobiological, and computational dimensions. Key research terms were categorized into four themes by analyzing diagnostic biomarker literature (Fig. 5B). This figure identifies three major research clusters, each exhibiting distinct research focus areas and characteristics. A detailed description of each cluster is as follows:

**Fig. 5.**
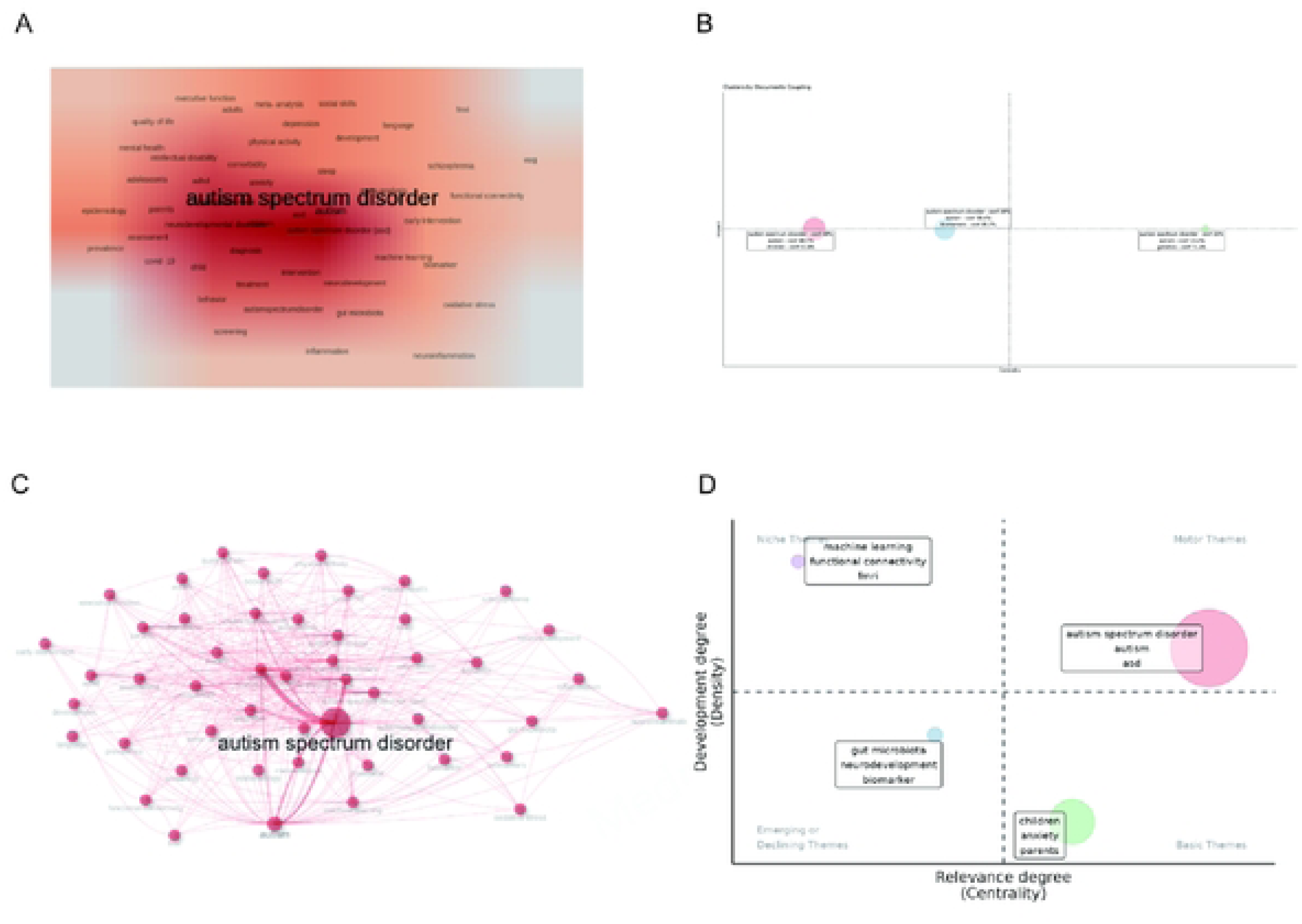
Comprehensive analysis of keywords and thematic structures in ASD research. (A) Keyword density over the past five years, with font size corresponding to search frequency. (B) Results of a coupling-clustering analysis applied to ASD literature. (C) Co-citation network of ASD-related topics; node size and connection thickness represent citation frequency and strength of association, respectively. (D) The thematicstructure of the ASD research field was examined using duster analysis and centrality metrics to identify research hotspots, core themes, and bridging terms that connect different thematic clusters.

Cluster 1 (108 publications): Primary label “autism spectrum disorder” (dominant in this cluster). The key component “Children’s confidence-57.1%” also plays a significant role. Centrality: 0.4498, indicating a highly central position in the network. Impact: 1.000, reflecting the substantial academic influence of the cluster. Clinically, this cluster underscores the importance of child-reported outcomes and self - confidence measures, which are often overlooked in biomarker-driven studies.

Cluster 2 (82 publications): Primary label “autism spectrum disorder”. The key component “gender-confidence 75%” is another major label. Centrality: 0.4034, slightly lower than Cluster 1 but still demonstrating strong network centrality. Impact: 0.500, signifying a high academic relevance. This cluster highlights the persistent emphasis on sex/gender differences, which have direct implications for tailored diagnostic and intervention strategies.

Cluster 3 (60 publications, the smallest cluster): Primary labels “autism spectrum disorder-confidence 22.7%,” “Biomarker-confidence 71.4%,” “Children-confidence 28.6%.” Centrality: 0.3698, relatively lower but still notable, given the cluster size. Impact: 0.25, indicating a consistent academic influence. The prominence of “biomarker” here reflects the field’s ongoing search for objective diagnostic tools, a key clinical gap that remains to be bridged.

Through co-citation clustering analysis, we identified three major ASD research clusters and elucidated their centrality and impact within the research network. These clusters reflect key trends and research focuses in ASD research from 2020 to 2025, underscoring the significance of gender/age differences and biomarkers in current research.

ASD-related exhibited strong associations with terms such as gut dysbiosis, early intervention, and neurodevelopment, indicating that the study of ASD, neurological development, and gut dysbiosis has increasingly become a recent hotspot (Fig. 5C). Thematic evolution mapping reveals a progressive shift in ASD research from foundational domains toward emerging interdisciplinary areas (Fig. 5D). Although a core cluster remains focused on definition, comorbidities, and interventions, two prominent emerging trajectories are identified: the gut-microbiota theme, which connects gastrointestinal ecology to neuroinflammation and biomarker research, and the machine learning theme, characterized by the application of computational models to neuroimaging and behavioral data. Research specifically focuses on children and integrates clinical, developmental, and psychosocial aspects.

### 3.5 Journal and bibliographic analysis

These seminal papers comprehensively investigated the etiology of ASD from multiple perspectives, including brain structure, genetics, psychology, gut microbiota, and animal models (Supplementary Table 1). Moreover, these publications appeared in prestigious journals with consistently high impact factors, such as Nature and New England Journal of Medicine (Supplementary Table 2). A clear inverse relationship exists between journal impact factors and citation counts in seminal ASD literature. Articles in high-impact-factor journals frequently demonstrate delayed citation accumulation despite their methodological advances, whereas seminal studies published in mid-tier journals often achieve exceptional, long-term citation dominance. Simultaneously, Chinese researchers remain significantly underrepresented in both high-IF and highly cited ASD publications. Therefore, future autism research should place greater emphasis on improving QoL and well-being of autistic children and their families, an approach widely endorsed in global guidelines but not yet sufficiently integrated into current research efforts, including those in China.

## 4. Discussion

### 4.1 Global Publication Trends and Persistent Research Gaps

The annual publication output in ASD research demonstrated a sustained upward trajectory over the 2020-2025 period, with annual volumes consistently exceeding 1,000 articles and a peak of 1710 publications in 2021 (Fig. 2A). This growth pattern aligns with the concurrently rising global prevalence estimates of ASD and the expanded diagnostic criteria introduced in the DSM-5[40,41], which have collectively increased both clinical demand and research investment over the past decade. The temporal peak observed in 2021 may partly reflect the scientific community’s accelerated response to the elevated mental health burden during the COVID-19 pandemic, when service disruptions and diagnostic delays were widely reported across multiple countries. The geographic distribution of research output remained highly concentrated, with the United States (2,768 publications), China (1,160), and the United Kingdom (615) comprising the top three contributing countries (Fig. 2B). This concentration of research activity underscores persistent geographic disparities in scientific capacity, which mirror the uneven distribution of clinical diagnostic and intervention resources worldwide-particularly in low- and middle-income countries where trained child psychiatrists and standardized screening tools remain scarce[42].

The keyword co-occurrence analysis identified three major thematic clusters in ASD research from 2020 to 2025: phenotyping (Cluster 1), gender/age differences (Cluster 2), and biomarkers (Cluster 3) (Fig. 5B). These clusters exhibit differential centrality and impact metrics: Cluster 1 (phenotyping) demonstrated the highest centrality (0.4498) and impact (1.000), whereas Cluster 3 (biomarkers) showed comparatively lower centrality (0.3698) and impact (0.25). This gradient may reflect the field’s stage of scientific maturation, where descriptive phenotyping-despite being methodologically established-continues to generate high-impact findings, whereas biomarker research, though actively pursued, has yet to produce clinically validated outputs of comparable citation impact[43]. The persistence of these clusters indicates that foundational etiological questions remain central to the field, and the continued emphasis on phenotyping and gender/age differences suggests that the clinical characterization of ASD remains an ongoing priority, particularly given the marked heterogeneity of the condition and the well-documented male-to-female ratio of 3-4:1[44].

However, the relatively low frequency of clinically oriented terms such as “quality of life,” and “early intervention” in the keyword landscape (Fig. 5A) suggests a translational gap between research outputs and pressing healthcare delivery needs. This gap is particularly salient given the rising prevalence of ASD and the corresponding demand for accessible, scalable service models in both high- and low-resource settings[45]. While basic science and etiological investigations have flourished, the translation of these findings into routine clinical practice-particularly in primary care settings, community-based services, and family-centered support systems-has not kept pace. Thematic evolution mapping further revealed two emerging trajectories, the gut-microbiota axis and machine learning applications (Fig. 5D), which although not yet forming independent major clusters during the study period, exhibited significantly increasing connectivity and growth momentum with core ASD themes[46]. This pattern suggests that these topics are nascent hotspots rather than fully established research paradigms[47]. The emergence of the gut-microbiota trajectory aligns with the broader scientific interest in the microbiome-neuroimmune interface, while the machine learning trajectory corresponds to the growing availability of multimodal datasets (neuroimaging, eye-tracking, behavioral) and computational tools. Given their relatively recent appearance in the publication record, these areas represent methodological shifts whose clinical utility and validity still require rigorous prospective evaluation[48].

### 4.2 Structural Asymmetries in Global Collaboration Networks

The collaboration network analysis revealed structural asymmetries in the global ASD research network. A notable pattern emerged from the comparison between Single Country Publication (SCP) and Multiple Country Publication (MCP) ratios: the United States, the United Kingdom, and Germany exhibited MCP ratios of 30%-40%, indicating strong international engagement, whereas China’s SCP ratio exceeded 50%, reflecting a predominantly domestic collaboration pattern (Fig. 3C). This pattern was further corroborated at the institutional level, where Chinese institutions showed sparse linkages with leading international centers, whereas North American and European institutions exhibited dense co-authorship networks (Fig. 4C). The contrast between the United States’ central position in the collaboration network-characterized by high betweenness centrality and extensive cross-border linkages-and China’s peripheral position, despite its substantial publication volume, represents a structural inefficiency in the global research enterprise[49].

Several factors may contribute to this disparity. First, language barriers in non-Anglophone research environments may limit the accessibility of Chinese research outputs to the broader international community and reduce opportunities for collaborative engagement[50]. Second, national funding mechanisms that prioritize domestic over international consortia may shape collaboration patterns, with Chinese research grants traditionally emphasizing home-grown investigations rather than multinational partnerships[51]. Third, data-sharing policies and privacy regulations may constrain cross-border collaborative research, particularly when involving human subjects and genomic data. These structural asymmetries have practical implications for the global ASD research enterprise, as they may limit the generalizability of findings derived from regionally confined datasets, reduce opportunities for methodological harmonization across diverse populations, and slow the translation of research discoveries into globally applicable clinical tools[52,53].

China’s position as the second-largest contributor in absolute publication volume, coupled with its comparatively limited international co-authorship, represents an underutilized collaborative potential[54]. Greater integration of Chinese research capacity into strategic international partnerships could accelerate progress in areas requiring large, diverse cohorts-particularly biomarker discovery, genetic epidemiology, and multi-omics studies, where data diversity is critical for generalizable findings. Moreover, international collaboration could facilitate the adoption of standardized diagnostic and intervention protocols, enhance the external validity of research findings, and promote capacity building in under-resourced regions . The relatively low international integration observed in the present analysis thus constitutes an opportunity for deliberate policy intervention to foster more inclusive and productive global partnerships in ASD research.

### 4.3 China’s Research Trajectory: Opportunities and Translational Challenges

The bibliometric evidence confirms that China has become the world’s second-largest contributor to ASD-related publications, with 1,160 publications over the study period. However, its research orientation, as reflected in keyword and institutional collaboration patterns, exhibits distinct characteristics. Chinese institutions demonstrated a stronger emphasis on molecular mechanisms (e.g., genetic variants, synaptic proteins) and early developmental phenotyping (e.g., high-risk infant cohorts), aligning with domestic funding priorities in basic neuroscience [55]. This mechanistic focus is consistent with China’s broader scientific strategy of investing in fundamental biomedical research, as reflected in national funding initiatives that prioritize basic science over translational or health services research. While this orientation has generated substantial progress in understanding the biological underpinnings of ASD, it has been pursued predominantly through domestic collaborations, as evidenced by the SCP ratio exceeding 50% [56]. This “domestic-first” pattern may limit the generalizability of findings across diverse genetic and environmental backgrounds, particularly given that ASD risk variants and their phenotypic expression vary across populations. Additionally, reduced exposure to internationally standardized diagnostic and intervention protocols may constrain the clinical applicability of research findings beyond the Chinese context.

From a healthcare delivery perspective, China faces substantial challenges that contextualize its research priorities: a large and geographically dispersed pediatric population, profound disparities in the distribution of child psychiatrists and developmental pediatricians between urban and rural areas, and limited reimbursement coverage for behavioral interventions. These systemic constraints create both a rationale and an opportunity for translating research capacity into practical, scalable solutions. Encouragingly, the emergence of “telemedicine” as a keyword in the bibliometric mapping (Fig. 5A), though still at low frequency, suggests nascent research interest in delivery-oriented solutions. Similarly, the growing attention to artificial intelligence-assisted screening tools and community-based intervention programs in recent Chinese publications reflects an increasing awareness of the need to bridge the gap between research discoveries and accessible clinical services [57]. However, these topics remain at the periphery of the Chinese ASD research landscape, indicating that substantial effort is still required to shift the research focus from mechanistic discovery to implementation science.

Nevertheless, a significant gap persists across both the Chinese and global literature: studies examining the quality of life (QoL) of autistic individuals and their families-encompassing employment outcomes, social participation, caregiver mental health, and educational inclusion-remained notably sparse in the Chinese literature, mirroring a global deficiency in this domain [58,59]. The Lancet Commission on the future of care and clinical research in autism has identified QoL as a core outcome measure, yet the bibliometric evidence suggests that this recommendation has not yet translated into commensurate research activity [16]. In the Chinese context, this gap is particularly concerning given the large population of autistic individuals and their families, the limited social support infrastructure, and the cultural factors that may influence caregiving burden and help-seeking behaviors. Closing this gap warrants national priority, requiring coordinated efforts from funding agencies, research institutions, and clinical service providers.

The three major clusters identified in the keyword co-occurrence analysis-phenotyping (Cluster 1), gender/age differences (Cluster 2), and biomarkers (Cluster 3)-represent active lines of inquiry, yet their varying centrality and impact metrics (Cluster 1: 0.4498, Cluster 2: 0.4034, Cluster 3: 0.3698) suggest differential academic influence. More critically, these clusters underscore persistent translational gaps: no clinically validated biomarker has emerged to date, and findings on gender differences have seldom been translated into sex-specific screening or intervention guidelines. To address these translational shortcomings, several actionable measures warrant consideration: routine screening for co-occurring conditions and systematic inclusion of QoL as a standard outcome measure should be incorporated into clinical practice; investment in telemedicine infrastructure and training programs for community-based ASD service providers-particularly in under-resourced areas-should be prioritized; and pragmatic trials conducted in real-world settings with diverse populations and patient-centered outcomes (adaptive behavior, family well-being) should be promoted [60]. The dual-perspective framework (global versus China) adopted in this study may also serve as a reference for other rapidly developing countries. By benchmarking against leading nations, emerging research producers can identify strategic niches-such as China’s mechanistic strengths-and leverage them to address local healthcare gaps, while simultaneously recognizing the importance of translational research and international collaboration for maximizing the societal impact of scientific investments.

## 5. Conclusions

This bibliometric analysis outlines the evolving global landscape of ASD research and demonstrates sustained growth in scientific output. Although the United States maintains its leading position, China has emerged as the second-largest contributor globally, with its institutions presenting greater emphasis on molecular mechanisms and early phenotyping, in line with national priorities. Important gaps remain regarding etiology, biomarker validation, and development of targeted interventions, underscoring the need for enhanced international collaboration. Future studies should prioritize standardized diagnostics, large-scale longitudinal cohorts with multi-omics data, biomarker-stratified clinical trials, and expansion of telemedicine. Improving the QoL among autistic individuals and their families must become a central goal. With its growing investment, China is well-positioned to contribute meaningfully to these challenges. To effectively translate research discoveries into clinical practice and scalable service models, however, it will be imperative to forge deeper global partnerships, enhance implementation science, and prioritize patient-centered outcome research that directly addresses the needs of autistic individuals and their families.

## Author Contributions

For research articles with several authors, a short paragraph specifying their individual contributions must be provided. The following statements should be used “Conceptualization, methodology, validation, formal analysis, resources,data curation and visualization are by Qianyi Zhong and Luping Chen; investigation by Yue Ji; writing-original draft preparation,review and editing by Qianyi Zhong; supervision by Fenglei Zhu; project administration and funding acquisition by Xiaobing Zou. All authors have read and agreed to the published version of the manuscript.”

## Institutional Review Board Statement

Not applicable. This study is a bibliometric analysis of publicly available published literature and did not involve human participants, animal subjects, or any personal data collection. Therefore, ethical approval was not required.

## Informed Consent Statement

Not applicable. This study did not involve human participants, patient data, or any identifiable personal information. Therefore, informed consent was not required.

## Data Availability Statement

The data presented in this study are derived from publicly available sources. The original publication records were retrieved from the Web of Science Core Collection (Science Citation Index Expanded) using the search strategy detailed in Section 2.1 (Materials and Methods). The specific search query, timeframe (January 2020-May 2025), and filtration criteria (document types: article, review, case report, clinical trial; removal of duplicates) are fully described in the manuscript to enable replication of the raw dataset. The processed data supporting the findings of this study, including the deduplicated records used for network visualizations and metrics, are available from the corresponding author upon reasonable request.

## Data Availability

The minimal dataset underlying the findings of this study is derived from the Web of Science Core Collection (Science Citation Index Expanded). The raw publication records were retrieved using the search strategy described in the Methods section (Section 2.1) of the manuscript, covering the period from January 2020 to May 2025. All analyzed data are publicly available from the Web of Science database however, access is subject to institutional subscription. The specific search query, inclusion criteria (document types: article, review, case report, clinical trial), and deduplication procedures are fully detailed in the manuscript to enable replication. The processed and deduplicated dataset used for bibliometric network analyses and visualizations is available from the corresponding author upon reasonable request.

## Acknowledgements

The authors thank all members of the Child Development Behavior Center at the Third Affiliated Hospital of Sun Yat-sen University for their support and collaboration. During the preparation of this manuscript, the authors used Deepseek for the purposes of language polishing and refining the expression of certain paragraphs. The authors have reviewed and edited the output and take full responsibility for the content of this publication.Conflicts of Interest: The authors declare no conflicts of interest.

---

**Supplementary Table 1.** Top10 most-cited references on ASD.

| Title | Date | Journal | IF | CC | Institution | Country |
| --- | --- | --- | --- | --- | --- | --- |
| Prevalence of Autism Spectrum Disorder Among Children Aged 8 Years - A<br>utism and Developmental Disabilities Monitoring Network, 11 Sites, United<br>States, 2016.<sup>[2]</sup> | 2020/<br>3/28 | MORBIDITY AND M<br>ORTALITY WEEKLY<br>REPORT. | 6.4 | 928 | National Center ,<br>CDC | United<br>States |
| Psychosocial and Behavioral Impact of COVID-19 in Autism Spectrum Diso<br>rder: An Online Parent Survey.<sup>[27]</sup> | 2020/<br>6/7 | BRAIN SCIENCES | 2.7 | 210 | University of<br>Verona | Italy |
| Prevalence and Characteristics of Autism Spectrum Disorder Among Childre<br>n Aged 8 Years - Autism and Developmental Disabilities Monitoring Networ<br>k, 11 Sites, United States, 2020.<sup>[28]</sup> | 2023/<br>3/23 | MORBIDITY AND M<br>ORTALITY WEEKLY<br>REPORT. | 6.4 | 147 | National<br>Center ,CDC | United<br>States |
| Autism spectrum disorder: definition, epidemiology, causes, and clinical eval<br>uation.<sup>[16]</sup> | 2020/<br>3/25 | TRANSLATIONAL P<br>EDIATRICS | 1.5 | 140 | Baylor College of<br>Medicine and Meyer<br>Center | United<br>States |
| Prevalence of comorbid psychiatric disorders among people with autism spec<br>trum disorder: An umbrella review of systematic reviews and meta-analyses.<sup>[</sup> | 2020/<br>3/24 | PSYCHIATRY RESE<br>ARCH | 4.2 | 115 | Texas A&M<br>University | United<br>States |
| Altered gut microbial profile is associated with abnormal metabolism activity<br>of Autism Spectrum Disorder<sup>[30]</sup> | 2020/<br>4/22 | GUT MICROBES | 12.2 | 114 | Nanjing Medical<br>University | China |
| Autism spectrum disorder.<sup>[3]</sup> | 2020/<br>1/18 | NATURE REVIEWS<br>DISEASE PRIMERS | 76.9 | 113 | University of<br>California | United<br>States |
| Composition of Gut Microbiota in Children with Autism Spectrum Disorder:<br>A Systematic Review and Meta-Analysis.<sup>[31]</sup> | 2020/<br>3/21 | NUTRIENTS | 4.8 | 104 | Universitat Rovira i<br>Virgili | Spain |
| Prevalence of Autism Spectrum Disorder in China: A Nationwide Multi-cent<br>er Population-based Study Among Children Aged 6 to 12Years.<sup>[24]</sup> | 2020/<br>7/2 | NEUROSCIENCE BU<br>LLETIN | 5.2 | 101 | Children's Hospital<br>of Fudan University | China |
| Oxidative Stress in Autism Spectrum Disorder.<sup>[32]</sup> | 2020/<br>2/7 | MOLECULAR NEUR<br>OBIOLOGY | 4.9 | 90 | Council for<br>Nutritional and<br>Environmental<br>Medicine | Norway |

CC,citation count.

**Supplementary Table 2.**
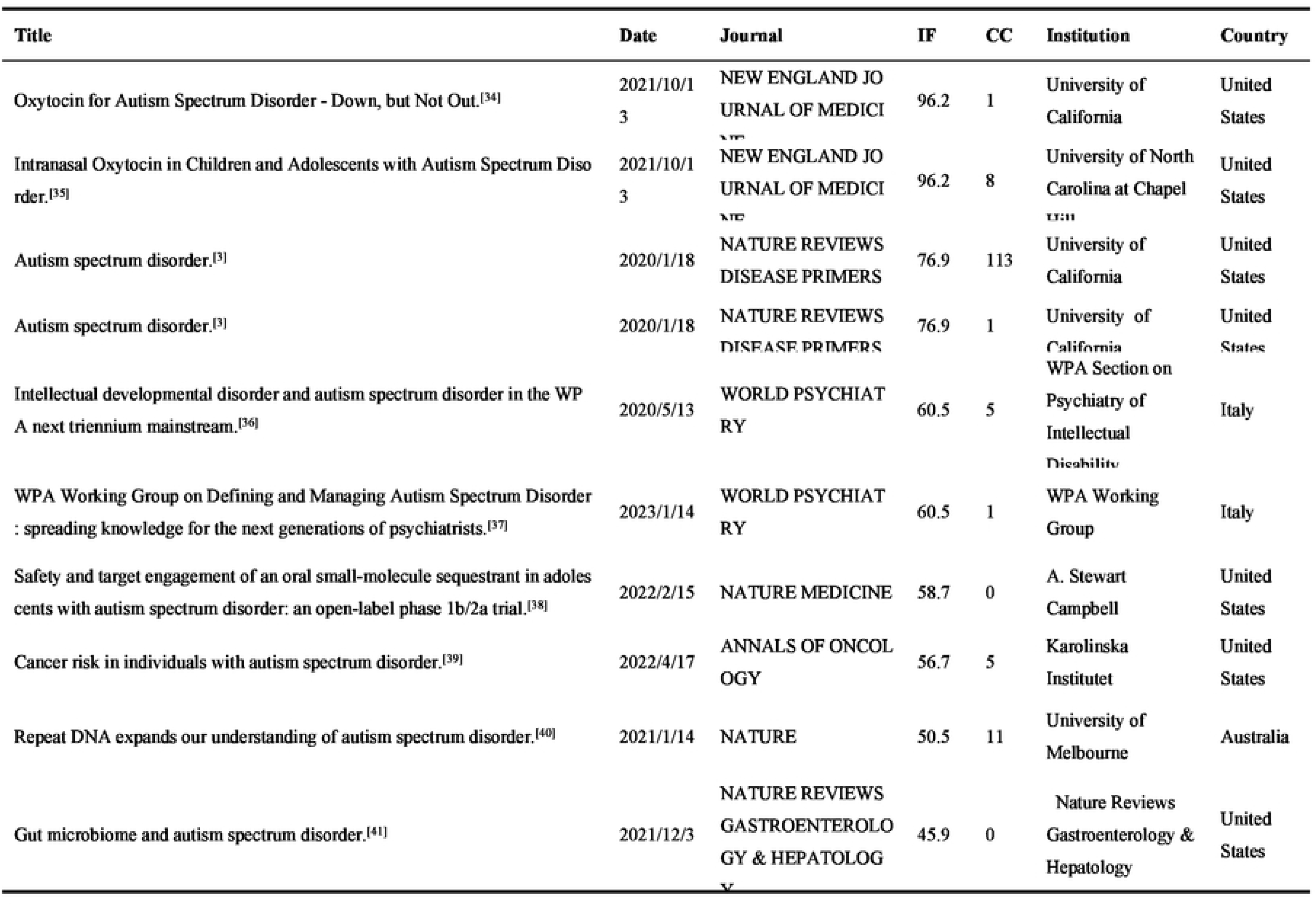
Top10 high-impact references on ASD.

